# Objective Sleep Quality in Diverse Older Adults: the Importance of Race and Ethnicity and Sex

**DOI:** 10.1101/2025.03.12.25323859

**Authors:** Clémence Cavaillès, Katie L. Stone, Yue Leng, Carrie Peltz, Kristine Yaffe

**Author notes:** Corresponding author: Clémence Cavaillès; 675 18^th^ Street, San Francisco, CA 94107.

## Abstract

**Background:** Research on sleep disparities across different sociodemographic groups is limited and often yields inconsistent findings. We aimed to examine differences in objective sleep measures by race and ethnicity, sex, and age within a diverse cohort of community-dwelling older adults.

**Methods:** We analyzed cross-sectional data from 838 participants aged ≥50 years in the Dormir Study (2020-2024). Sleep metrics, including sleep duration, sleep efficiency, wake after sleep onset (WASO), and sleep fragmentation index (SFI), were derived from 7-day wrist actigraphy. Race and ethnicity (Black; Mexican American [MA]; Non-Hispanic White [NHW]), sex, and age (<65; ≥65 years) were self-reported. We compared sleep metrics across sociodemographic groups and assessed their multivariable associations using linear, logistic, and multinomial regression models.

**Results:** We studied 190 (22.7%) Black, 282 (33.6%) MA, and 366 (43.7%) NHW Dormir participants, with a mean age of 66.7 ±8.4 years, and 64.8% women. Compared to NHW participants, Black and MA participants had shorter mean sleep duration (Black: 7.1 ±1.2 hours; MA: 7.1 ±1.1 hours; NHW: 7.5 ±1.1 hours; p<0.0001), lower median sleep efficiency (Black: 87.2%; MA: 87.8%; NHW: 90.6%; p<0.0001), longer median WASO (Black: 61.2 minutes; MA: 56.7 minutes; NHW: 44.4 minutes; p<0.0001), and higher mean SFI (Black: 32.0 ±11.0%; MA: 27.3 ±9.7%; NHW: 24.0 ±9.0%; p<0.0001). Compared to men, women had longer mean sleep duration (women: 7.4 ±1.1 hours; men: 7.1 ±1.2 hours; p=0.0004) and lower mean SFI (women: 25.9 ±8.8%; men: 28.9 ±12.1%; p=0.0001). Older participants had longer mean sleep duration (old: 7.4 ±1.1 hours; young: 7.1 ±1.1 hours; p<0.0001), higher median sleep efficiency (old: 89.8%; young: 87.7%; p<0.0001), shorter median WASO (old: 48.5 minutes; young: 56.8 minutes; p<0.0001), and lower mean SFI (old: 26.1 ±10.2%; young: 28.1 ±10.2%; p=0.007). After adjusting for socioeconomic and behavioral factors, comorbidities, and sleep medications, findings were consistent except for age group comparisons in which differences were no longer significant.

**Conclusions:** Our findings demonstrate significant variations in objective sleep measures across sociodemographic groups, with non-White participants and men experiencing poorer sleep quality. These disparities may contribute to health inequalities, emphasizing the need for targeted interventions to support at-risk populations.

## Background

Poor sleep quality is common in the general population and has been associated with a wide range of physical and mental health issues, including metabolic and cardiovascular disease, cognitive impairment, depression, and an increased mortality risk [1–5]. Understanding how sleep measures vary across sociodemographic groups and identifying populations that experience worse sleep are critical for informing targeted public health interventions. These efforts may help mitigate the burden of poor sleep and address broader health disparities. This is particularly important for older adults, who are already at greater risk for adverse health outcomes and for whom poor sleep may exacerbate or accelerate the onset of age-related diseases.

Race and ethnicity are a key sociodemographic factor that may highlight sleep disparities [6,7]. A few studies have investigated racial and ethnic minorities, such as Black adults, and reported worse sleep outcomes compared to White adults [8–10]. However, research examining sleep across diverse racial and ethnic populations remains limited. Most studies have primarily focused on sleep differences between White and Black individuals, leaving other minority groups, such as Hispanics, relatively understudied [11]. Moreover, most prior studies relied on self-reported sleep data, which is prone to bias and often overlooks sleep features that require objective measurement for good accuracy like sleep fragmentation [11]. Actigraphy, an objective, non-invasive, and cost-effective method for sleep assessment, offers a valuable alternative.

Furthermore, sex differences may also play an important role in sleep health [12]; but the relationship between sleep measures and sex is inconsistent and appears to vary according to the method of sleep assessment. Studies based on self-reported sleep data have generally indicated that women reported more sleep complaints, insomnia symptoms, and experienced poorer sleep quality and shorter sleep duration than men [13–17]. In contrast, studies using objective sleep measures have suggested that women may have better objective sleep quality, less fragmented sleep, and longer sleep duration compared to men [13,15,16,18,19]. Previous research has also shown that sleep quality declines with age, as evidenced by longer periods of wakefulness after sleep onset, more frequent nighttime awakenings, reduced sleep efficiency, decreased nighttime sleep duration, and higher frequency of sleep disorders [13,18,20–22]. However, the combined effects of sex, age, and race and ethnicity in relation to sleep differences remain unexplored.

To address these gaps, we examined differences in objective sleep measures by race and ethnicity, sex, and age groups, as well as their combinations, using actigraphy-derived data from a recently assembled large, diverse cohort of community-dwelling older adults, including Non-Hispanic White (NHW) participants and nearly 60% of non-White (Black and Mexican American [MA]) individuals. We also assessed whether sleep disparities among diverse populations are independent of socioeconomic status and health factors.

## Methods

### Study Design

We conducted a sleep study ancillary to the Health & Aging Brain Study – Health Disparities (HABS-HD), the Dormir Study. Briefly, HABS-HD is a longitudinal, community-based study of cognitive aging among Black, MA, and NHW participants, conducted by the University of North Texas Health Science Center [23]. Subjects were enrolled in the study baseline visit using a community-based participatory research approach for reaching underserved and racial and ethnic minority populations, leading to a representative sample of the larger community [23].

Inclusion criteria included age 50 and older, self-reported race and ethnicity of Black, MA, or NHW, fluency in English or Spanish; detailed HABS-HD protocol has been described previously [23].

The Dormir Study was conducted between 2020 and 2024 and invited eligible HABS-HD participants to undergo a comprehensive sleep examination, including actigraphy and a sleep diary. Participants who had agreed to be re-contacted from the HABS-HD cohort were invited to complete a brief pre-screening questionnaire if they were interested. Those with permanent pacemakers and those currently using a continuous positive airway pressure machine were excluded. Our study included 838 participants with complete sociodemographic data as of May 2024 and valid actigraphy data (defined as wearing the watch for at least four nights and maintaining a sleep diary). All study participants provided written informed consent, and the Dormir Study was approved by the University of North Texas Health Science Center Institutional and University of California San Francisco Review Boards.

### Actigraphy

Actigraphy data was collected using the GT3XActiGraph Link device (ActiGraph, Inc.). Participants were instructed to wear this watch-like device on their nondominant wrist continuously (except for bathing) for a minimum of 4 and up to 7 consecutive days and nights. Data were scored in 60-second epochs, and the Cole-Kripke sleep scoring algorithm was applied using ActiLife 6 software to determine “sleep” and “wake” periods [24]. Trained scorers edited the data using participants’ sleep diaries to identify (i) in-bed and out-of-bed intervals; and (ii) non-wear times, which were removed, by using either the declared off-wrist sensor or the Troiano non-wear algorithm [25,26]. The ActiLife software option was used to set all non-wear periods within the in-bed interval as sleep for calculations in the nightly in-bed interval summaries. Sleep measures were averaged across all days and nights of recording.

We examined four measures of nighttime sleep: sleep duration (hours), sleep efficiency (ratio of sleep duration to time spend in bed, multiplied by 100), wake after sleep onset (WASO; minutes of wake during the sleep interval), and sleep fragmentation index (SFI; sum of the percentage of the sleep period spent moving and the percentage of immobile periods ≤1 minute). We examined these measures continuously and categorically considering commonly used cut-offs for sleep duration (<6, 6-8, >8 hours) and sleep efficiency (<85, ≥85%), and WASO and SFI tertile.

### Covariates

Sociodemographic and health characteristics, including age, sex, race and ethnicity, education, retirement status, household income, current smoking status, and medical history of hypertension, diabetes, stroke, heart attack, and depression, were self-reported through clinical interviews. Body mass index (BMI) was calculated and physical activity measured using the Rapid Assessment of Physical Activity (RAPA) questionnaire [27]. Cognitive status (normal control; cognitive impairment [mild cognitive impairment and dementia]) was determined through an algorithmic decision tree and consensus review [23]. Sleep medication use was self-reported from the Pittsburgh Sleep Quality Index [28]. In a subsample (n=784), sleep apnea was assessed using the WatchPAT 200 (Itamar Medical Ltd.) [29], with a respiratory event index (REI) ≥15 indicating moderate to severe sleep apnea (see eMethods).

### Statistical Analysis

Participants’ characteristics were compared by race and ethnicity using chi-squared tests for categorical variables, ANOVA for normally distributed continuous variables, and Kruskal-Wallis tests for skewed continuous variables. Sleep measures were compared by sex and age groups using chi-squared, t-tests, and Mann-Whitney U tests. Sleep comparisons by sex were further stratified by race and ethnicity (six groups) and analyzed similarly. We presented frequencies for categorical variables, mean (± standard deviation) for normally distributed continuous variables, and median (interquartile) otherwise.

We examined the multivariable associations between sleep measures and sociodemographic groups using linear, logistic, and multinomial logistic regressions. Models were adjusted for race and ethnicity, sex, age, education, income, cognitive status, retirement status, smoking, physical activity, BMI, sleep medications, and history of hypertension, diabetes, stroke, heart attack, and depression. Non-normally distributed variables were transformed (cube and log transformations) with results back-transformed to their original scale. Adjusted means (95% confidence intervals [CI]) were reported for continuous variables and odds ratios (95% CI) for categorical variables. Sensitivity analyses included False Discovery Rate (FDR) corrections for multiple testing and further adjustment for income (n=787; due to missing data) and sleep apnea (n=784).

Furthermore, we used a stepwise approach to assess the impact of covariates on the association between sleep measures and age groups. Notable changes in effect size, loss of significance, or substantial improvement in adjusted R^2^ indicated that the covariate explained meaningful variation in the outcome. All statistical tests were two-sided, and analyses were performed using R version 4.4.0.

## Results

The 838 participants had a mean age of 66.7±8.4 years and 64.8% were women, 190 (22.7%) were Black, 282 (33.6%) were MA, and 366 (43.7%) were NHW. Compared to NHW participants, Black and MA participants were more likely to be women, younger, employed, smokers, and to have lower incomes. They also had higher BMI, as well as higher rates of cognitive impairment, history of hypertension and diabetes, and sleep medication use.

Additionally, MA participants had fewer years of education and were less physically active than NHW participants (Table 1).

**Table 1.** Description of covariates by race and ethnicity (n=838).

|  | Total sample<br>(n=838) |  | Black<br>(n=190) | MA<br>(n=282) | NHW<br>(n=366) | *P*<br>value<sup>b</sup> |
| --- | --- | --- | --- | --- | --- | --- |
|  | Missing<br>data, No. | No. (%) or<br>mean (±SD) | No. (%) or<br>mean (±SD) | No. (%) or<br>mean (±SD) | No. (%) or<br>mean (±SD) |  |
| Sex, *women* | 0 | 543 (64.8) | 131 (68.9) | 198 (70.2) | 214 (58.5) | 0.003 |
| Age, years | 0 | 66.7 (±8.4) | 63.5 (±7.5) | 64.5 (±7.9) | 70.1 (±8.1) | <.0001 |
| Age, ≥65 years | 0 | 492 (58.7) | 83 (43.7) | 136 (48.2) | 273 (74.6) | <.0001 |
| Education, years | 0 | 13.8 (±4.0) | 15.0 (±2.6) | 10.5 (±4.2) | 15.7 (±2.7) | <.0001 |
| Retirement status | 2 | 507 (60.6) | 100 (52.6) | 136 (48.4) | 271 (74.2) | <.0001 |
| Income group | 38 |  |  |  |  | <.0001 |
| <25,000 |  | 215 (26.9) | 58 (34.1) | 117 (44.2) | 40 (11.4) |  |
| [25,000-50,000[ |  | 193 (24.1) | 37 (20.0) | 81 (30.6) | 75 (21.4) |  |
| [50,000-75,000[ |  | 124 (15.5) | 27 (14.6) | 36 (13.6) | 61 (17.4) |  |
| ≥75,000 |  | 268 (33.5) | 63 (34.1) | 31 (11.7) | 174 (49.7) |  |
| Cognitive impairment | 0 | 188 (22.4) | 59 (31.1) | 72 (25.5) | 57 (15.6) | <.0001 |
| Smoke currently | 0 |  |  |  |  |  |
| RAPA score<sup>a</sup> | 0 | 4 (3-6) | 4 (3-6) | 4 (3-4.8) | 4 (3-6) | 0.0004 |
| BMI, kg/m<sup>2</sup><sup>a</sup> | 10 | 29.8 (26.1,34.7) | 32.1 (27.3,37.5) | 29.8 (26.9,35.3) | 29.2 (24.7,32.9) | <.0001 |
| Sleep medications | 0 | 307 (36.6) | 67 (35.3) | 85 (30.1) | 155 (42.3) | 0.005 |
| History of hypertension | 0 | 537 (64.1) | 149 (78.4) | 173 (61.3) | 215 (58.7) | <.0001 |
| History of diabetes | 0 | 210 (25.1) | 57 (30.0) | 98 (34.8) | 55 (15.0) | <.0001 |
| History of stroke | 0 | 42 (5.0) | 12 (6.3) | 7 (2.5) | 23 (6.3) | 0.06 |
| History of heart attack | 2 | 33 (3.9) | 10 (5.3) | 11 (3.9) | 12 (3.3) | 0.53 |
| History of depression | 0 | 290 (34.6) | 58 (30.5) | 108 (38.3) | 124 (33.9) | 0.20 |
| REI, ≥15 | 54 | 399 (50.9) | 90 (53.6) | 141 (52.6) | 168 (48.3) | 0.42 |

Abbreviations: BMI, body mass index; MA, Mexican American; NHW, Non-Hispanic White; RAPA, rapid assessment of physical activity; REI, respiratory event index; SD, standard deviation
<sup>a</sup> median (interquartile range)
<sup>b</sup> Anova test was used for continuous variables with a normal distribution, Kruskal-Wallis test for continuous variables without a normal distribution and, Chi-square test for categorical variables

### Race and ethnic differences in sleep

The average sleep duration among all participants was 7.3±1.1 hours, with a median sleep efficiency of 89.0%, a median WASO of 51.1 minutes, and a mean SFI of 26.9±10.2%.

Compared to NHW participants, Black and MA participants had shorter mean sleep duration (Black: 7.1±1.2 hours; MA: 7.1±1.1 hours; NHW: 7.5±1.1 hours; p<0.0001), lower median sleep efficiency (Black: 87.2%; MA: 87.8%; NHW: 90.6%; p<0.0001), longer median WASO (Black: 61.2 minutes; MA: 56.7 minutes; NHW: 44.4 minutes; p<0.0001), and higher mean SFI (Black: 32.0±11.0%; MA: 27.3±9.7%; NHW: 24.0±9.0%; p<0.0001) (Table 2). The prevalence of short sleep duration, poor sleep efficiency, long WASO, and high SFI were highest among Black participants, followed by MA participants, and then NHW participants (Table 2). These racial and ethnic differences remained statistically significant for all sleep measures, except for the prevalence of short sleep duration and high SFI in MA participants after adjusting for potential confounders (Table 3, eTable 1). Moreover, the differences remained significant after FDR correction and further adjustment for income and sleep apnea (eTables 2-5).

**Table 2.** Objective sleep measures by sociodemographic groups (n=838).

|  | Black<br>(n=190) | MA<br>(n=282) | NHW<br>(n=366) |  | Men<br>(n=295) | Women<br>(n=543) |  | <65 years<br>(n=346) | ≥65 years<br>(n=492) |  |
| --- | --- | --- | --- | --- | --- | --- | --- | --- | --- | --- |
|  | No. (%) or<br>mean (±SD) | No. (%) or<br>mean (±SD) | No. (%) or<br>mean (±SD) | *P*<br>value<sup>b</sup> | No. (%) or<br>mean (±SD) | No. (%) or<br>mean (±SD) | *P*<br>value<sup>c</sup> | No. (%) or<br>mean (±SD) | No. (%) or<br>mean (±SD) | *P*<br>value<sup>c</sup> |
| **Sleep duration, hour** |  |  |  | <.0001* |  |  | 0.002* |  |  | 0.002* |
| <6 | 38 (20.0) | 42 (14.9) | 23 (6.3) |  | 51 (17.3) | 52 (9.6) |  | 54 (15.6) | 49 (10.0) |  |
| [6-8] | 115 (60.5) | 186 (66.0) | 238 (65.0) |  | 187 (63.4) | 352 (64.8) |  | 229 (66.2) | 310 (63.0) |  |
| >8 | 37 (19.5) | 54 (19.1) | 105 (28.7) |  | 57 (19.3) | 139 (25.6) |  | 63 (18.2) | 133 (27.0) |  |
| *Continuous* | 7.1 (±1.2) | 7.1 (±1.1) | 7.5 (±1.1) | <.0001* | 7.1 (±1.2) | 7.4 (±1.1) | 0.0004* | 7.1 (±1.1) | 7.4 (±1.1) | <.0001* |
| **Sleep efficiency, %** |  |  |  | <.0001* |  |  | 0.09 |  |  | 0.001* |
| ≥85 | 124 (65.3) | 192 (68.1) | 318 (86.9) |  | 213 (72.2) | 421 (77.5) |  | 242 (69.9) | 392 (79.7) |  |
| <85 | 66 (34.7) | 90 (31.9) | 48 (13.1) |  | 82 (27.8) | 122 (22.5) |  | 104 (30.1) | 100 (20.3) |  |
| *Continuous*<sup>a</sup> | 87.2 (83.3,90.0) | 87.8 (83.6,90.8) | 90.6 (87.2,93.0) | <.0001* | 89.0 (84.1,92.3) | 89.0 (85.3,91.8) | 0.61 | 87.7 (84.2,90.9) | 89.8 (86.3,92.4) | <.0001* |
| **WASO, min** |  |  |  | <.0001* |  |  | 0.12 |  |  | 0.001* |
| <41.7 | 41 (21.6) | 68 (24.1) | 170 (46.4) |  | 110 (37.3) | 169 (31.1) |  | 93 (26.9) | 186 (37.8) |  |
| [41.7-63.5] | 64 (33.7) | 99 (35.1) | 117 (32.0) |  | 87 (29.5) | 193 (35.5) |  | 117 (33.8) | 163 (33.1) |  |
| >63.5 | 85 (44.7) | 115 (40.8) | 79 (21.6) |  | 98 (33.2) | 181 (33.3) |  | 136 (39.3) | 143 (29.1) |  |
| *Continuous*<sup>a</sup> | 61.2 (44.4,79.8) | 56.7 (42.2,78.0) | 44.4 (30.6,60.5) | <.0001* | 48.6 (34.3,73.4) | 52.7 (37.3,68.5) | 0.51 | 56.8 (40.4,75.6) | 48.5 (33.3,66.9) | <.0001* |
| **SFI, %** |  |  |  | <.0001* |  |  | 0.006* |  |  | 0.004* |
| <22.1 | 28 (14.7) | 88 (31.2) | 164 (44.8) |  | 92 (31.2) | 188 (34.6) |  | 94 (27.2) | 186 (37.8) |  |
| [22.1-29.9] | 62 (32.6) | 97 (34.4) | 119 (32.5) |  | 84 (28.5) | 194 (35.7) |  | 120 (34.7) | 158 (32.1) |  |
| >29.9 | 100 (52.6) | 97 (34.4) | 83 (22.7) |  | 119 (40.3) | 161 (29.7) |  | 132 (38.2) | 148 (30.1) |  |
| *Continuous* | 32.0 (±11.0) | 27.3 (±9.7) | 24.0 (±9.0) | <.0001* | 28.9 (±12.1) | 25.9 (±8.8) | 0.0001* | 28.1 (±10.2) | 26.1 (±10.2) | 0.007* |

Abbreviations: CI, confidence interval; MA, Mexican American; NHW, Non-Hispanic White; SD, standard deviation; SFI, sleep fragmentation index; WASO, wake after sleep onset
<sup>a</sup> median (interquartile range)
<sup>b</sup> Anova test was used for continuous variables with a normal distribution, Kruskal-Wallis test for continuous variables without a normal distribution and, Chi-square test for categorical variables
<sup>c</sup> T-test was used for continuous variables with a normal distribution, Mann-Whitney U test for continuous variables without a normal distribution and, Chi-square test for categorical variables
\* results are significant after false discovery rate correction

**Table 3.**
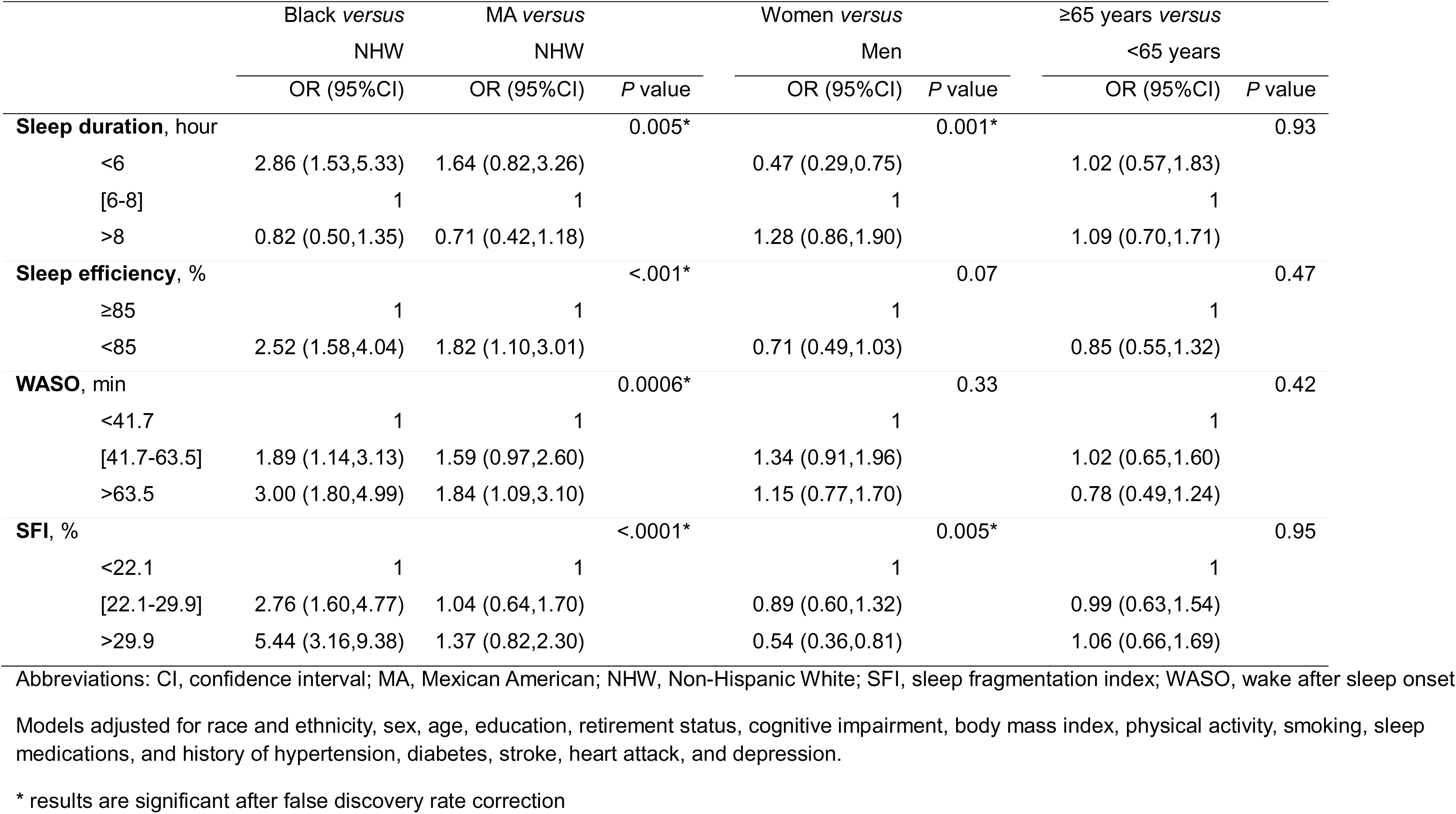
Multivariable-adjusted associations between objective sleep measures and sociodemographic groups (n=824).

|  | Black *versus*<br>NHW | MA *versus*<br>NHW | *P* value | Women *versus*<br>Men | *P* value | ≥65 years *versus*<br><65 years | *P* value |
| --- | --- | --- | --- | --- | --- | --- | --- |
|  | OR (95%CI) | OR (95%CI) |  | OR (95%CI) |  | OR (95%CI) |  |
| **Sleep duration, hour** |  |  | 0.005* |  | 0.001* |  | 0.93 |
| <6 | 2.86 (1.53,5.33) | 1.64 (0.82,3.26) |  | 0.47 (0.29,0.75) |  | 1.02 (0.57,1.83) |  |
| [6-8] | 1 | 1 |  | 1 |  | 1 |  |
| >8 | 0.82 (0.50,1.35) | 0.71 (0.42,1.18) |  | 1.28 (0.86,1.90) |  | 1.09 (0.70,1.71) |  |
| **Sleep efficiency, %** |  |  | <.001* |  | 0.07 |  | 0.47 |
| ≥85 | 1 | 1 |  | 1 |  | 1 |  |
| <85 | 2.52 (1.58,4.04) | 1.82 (1.10,3.01) |  | 0.71 (0.49,1.03) |  | 0.85 (0.55,1.32) |  |
| **WASO, min** |  |  | 0.0006* |  | 0.33 |  | 0.42 |
| <41.7 | 1 | 1 |  | 1 |  | 1 |  |
| [41.7-63.5] | 1.89 (1.14,3.13) | 1.59 (0.97,2.60) |  | 1.34 (0.91,1.96) |  | 1.02 (0.65,1.60) |  |
| >63.5 | 3.00 (1.80,4.99) | 1.84 (1.09,3.10) |  | 1.15 (0.77,1.70) |  | 0.78 (0.49,1.24) |  |
| **SFI, %** |  |  | <.0001* |  | 0.005* |  | 0.95 |
| <22.1 | 1 | 1 |  | 1 |  | 1 |  |
| [22.1-29.9] | 2.76 (1.60,4.77) | 1.04 (0.64,1.70) |  | 0.89 (0.60,1.32) |  | 0.99 (0.63,1.54) |  |
| >29.9 | 5.44 (3.16,9.38) | 1.37 (0.82,2.30) |  | 0.54 (0.36,0.81) |  | 1.06 (0.66,1.69) |  |

Abbreviations: CI, confidence interval; MA, Mexican American; NHW, Non-Hispanic White; SFI, sleep fragmentation index; WASO, wake after sleep onset
Models adjusted for race and ethnicity, sex, age, education, retirement status, cognitive impairment, body mass index, physical activity, smoking, sleep medications, and history of hypertension, diabetes, stroke, heart attack, and depression.
\* results are significant after false discovery rate correction

### Sex differences in sleep

Sleep measures differed by sex (Table 2). Compared to men, women had longer mean sleep duration (7.1±1.2 *vs.* 7.4±1.1 hours; p=0.0004) and lower mean SFI (28.9±12.1 *vs.* 25.9±8.8%; p=0.0001). Women also showed lower prevalence of short sleep duration, high SFI, and higher prevalence of long sleep duration; with a tendency for higher rates of good sleep efficiency (p=0.09) and short WASO (p=0.12). In the multivariable model, women displayed lower odds of short sleep duration and high SFI (highest tertile) than men (Table 3). The adjusted mean sleep duration and SFI were 7.0 hours and 32.4% for men and 7.3 hours and 29.1% for women, respectively (eTable 6). Sensitivity analyses displayed comparable findings (eTables 2, 4, 7, 8).

Figure 1 illustrates the sleep measures by sex and race and ethnicity. Among the six groups, NHW women exhibited the longest mean sleep duration, highest median sleep efficiency, and lowest median SFI, alongside the highest prevalence of long sleep duration, good sleep efficiency and lowest prevalence of long WASO and high SFI (eTable 9). NHW men had the shortest median WASO. In contrast, Black men exhibited the shortest mean sleep duration, lowest median sleep efficiency, longest median WASO, and highest mean SFI. They also had the highest prevalence of short sleep duration, poor sleep efficiency, long WASO, and high SFI (eTable 9).

**Figure 1.**
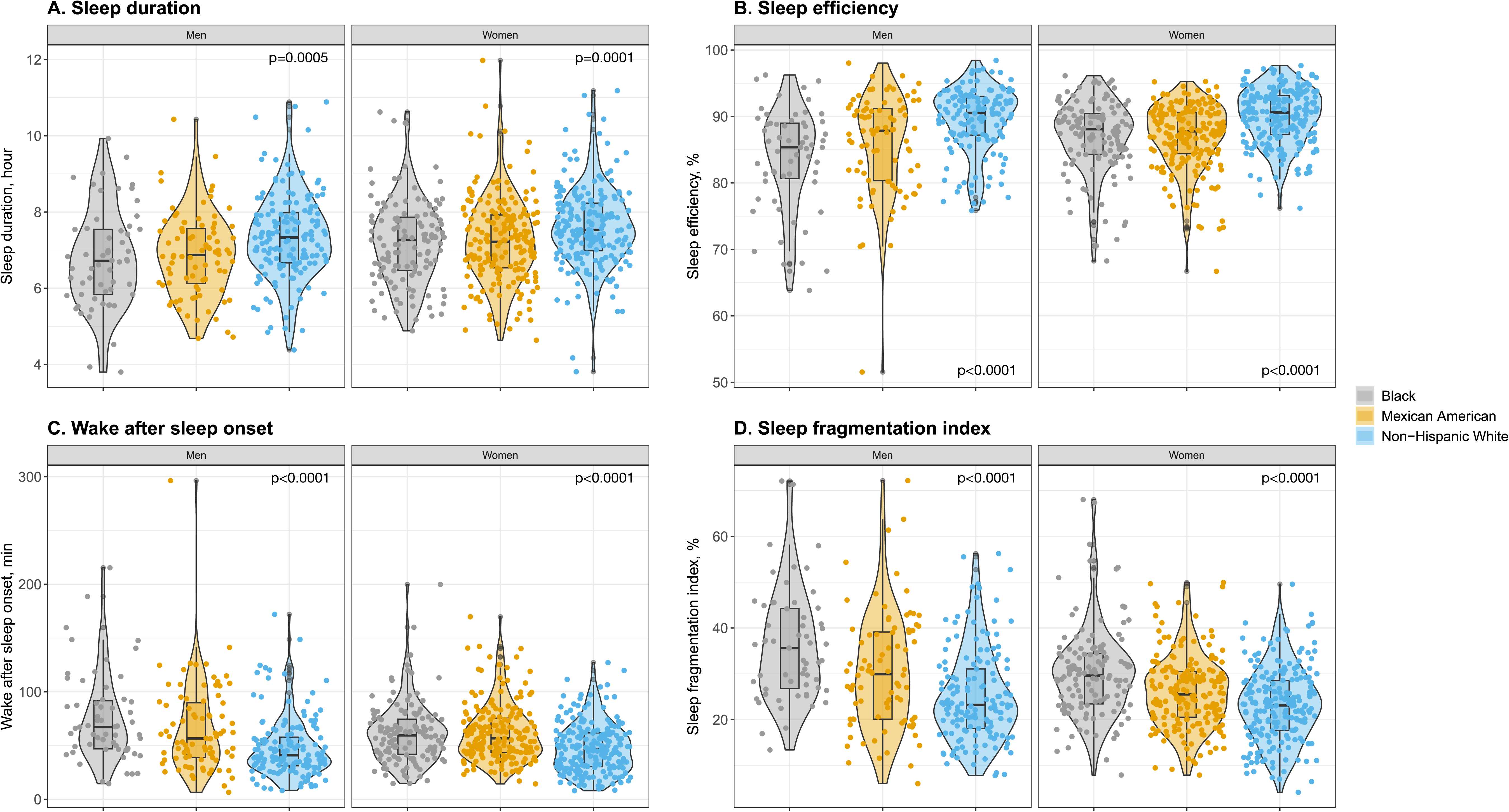
Objective sleep measures by race and ethnicity and sex. Anova test was used for continuous variables with a normal distribution and Kruskal-Wallis test for continuous variables without a normal distribution.

### Age differences in sleep

Compared to younger participants (<65 years), older participants (≥65 years) had longer mean sleep duration (7.1±1.1 *vs.* 7.4±1.1 hours; p<0.0001), higher median sleep efficiency (87.7 *vs.* 89.8%; p<0.0001), shorter median WASO (56.8 *vs.* 48.5 minutes; p<0.0001), and lower mean SFI (28.1±10.2 *vs.* 26.1±10.2%; p=0.007). They also exhibited higher prevalence of long sleep duration and lower prevalence of short sleep duration, poor sleep efficiency, long WASO, and high SFI (Table 2). However, after controlling for potential confounders, sleep measures no longer differed between the two groups (Table 3, eTable 6). The main covariates driving the association were race and ethnicity and retirement status (eFigures 1-4). Results remained consistent in the sensitivity analyses (eTables 2, 4, 7, 8).

## Discussion

In a diverse sample of community-dwelling older adults, we identified disparities in actigraphy-derived sleep measures across race and ethnicity and sex. Compared to NHW adults, Black and MA participants had shorter sleep duration and worse sleep quality, with lower sleep efficiency and more fragmented sleep. Additionally, women experienced better sleep than men, with longer sleep duration and lower SFI. With further stratification, White women generally demonstrated the most favorable sleep outcomes, while Black men exhibited the poorest. After adjusting for potential confounders, we did not find sleep differences between age groups.

Overall, these findings indicate that objective poor sleep is more frequent among non-White participants and among men, potentially contributing to broader health disparities.

Our findings align with a few prior studies, suggesting that non-White individuals have worse sleep outcomes than White adults [6,7,11]. Our results are notable for including several race and ethnic groups and investigating sex and age groups as well. These are particularly important as Hispanics remain underrepresented in sleep research, despite being the largest racial and ethnic minority group in the US [30]. Given that the Hispanic population aged 65 and older is projected to triple by 2050 [31], addressing sleep disparities in this group is a growing public health priority. Only a few studies have used actigraphy to investigate sleep in several racial and ethnic minority groups [8,9,32]. Moreover, these studies typically involve small sample sizes with disproportionately fewer non-White participants and fail to adjust for major confounders like socioeconomic status, depression, or sleep medications. For example, Chen et al., in the Multi-Ethnic Study of Atherosclerosis, reported higher odds of short sleep duration among Black, Chinese, and Hispanic participants compared to NHW adults and that Black participants had poorer sleep efficiency than NHWs, though no significant differences were observed for Chinese and Hispanic participants [8]. However, this study did not account for education, income, or important health conditions. In contrast, our study controlled for a broad range of socioeconomic, behavioral, and comorbidity variables, and yet significant disparities persisted. The causes of these disparities are likely multifactorial and may include stress, environmental factors (e.g., housing, neighborhood environment, pollution), psychological or medical factors [33–35]. Structural racism and discrimination may also contribute to these disparities, in part by limiting access to quality healthcare and increasing psychological distress [33–35]. Future studies should explore more deeply the role of these potential drivers in diverse populations.

We also found that women had objectively longer sleep duration and lower SFI than men, consistent with some previous studies [13,15,16,18,19]. Although the mechanisms underlying these sex differences are not well understood, several hypotheses have been suggested. One suggests that men tend to have unhealthier lifestyles than women [36,37], possibly leading to poorer sleep quality. However, our findings indicate that sex differences in sleep persist even after accounting for numerous behavioral and comorbidity factors, suggesting that lifestyle may not be the primary driver of these differences. Hormonal factors may also play a role. In women, progesterone has been shown to improve sleep onset latency and sleep continuity [38–40], while estrogen would enhance rapid-eye movement sleep [38]. In contrast, testosterone levels, which decline with age in men, have been linked to poorer sleep quality [39,41]. Additionally, sleep architecture differences may contribute to these disparities, as women generally have more slow-wave sleep, the deepest and most restorative sleep stage [19,20,42]. Interestingly, although women had lower fragmented sleep based on the SFI, no differences were observed for WASO, another marker of sleep fragmentation. SFI captures the frequency of sleep interruptions and movements, including brief arousals, whereas WASO represents the total time spent awake after falling asleep. Thus, our results suggest that men may experience more frequent but shorter sleep disruptions than women.

When examining the interaction between race and ethnicity and sex, striking patterns emerged. NHW women demonstrated the most favorable sleep patterns, while Black men exhibited the least favorable sleep measures. Both MA men and women and Black women generally showed intermediate sleep measures. These findings are consistent with a prior study of middle-aged adults, showing longer sleep duration and higher sleep efficiency among White women, followed by White men, Black women, and finally Black men [43]. Our study greatly adds to these observations to older adults and includes MA participants, providing a broader understanding of sleep disparities across diverse populations.

Contrary to previous studies [13,20,44,45], our unadjusted age-stratified analysis revealed that older participants had better sleep compared to younger participants. This discrepancy could stem from our sample’s relatively narrow age range, potentially limiting the ability to capture broader age-related trends. It is also possible that our older participants represent a healthier subset with subsequent better sleep (eTable 10). Another explanation is that, as most prior studies have been conducted in White populations, they may have missed important disparities driven by race and ethnicity. Indeed, our multivariate analysis showed that the sleep differences were no longer significant after adjusting for the potential confounders, and particularly race and ethnicity and retirement status. This suggests that sleep disparities are more strongly influenced by a combination of racial and ethnic background and socioeconomic factors than by aging alone. Therefore, sleep improvement strategies should target specific challenges like work stress or systemic barriers.

Taken together, our findings highlight that race and ethnicity and sex are key contributors to sleep disparities, independent of socioeconomic and health factors, with age being of lesser importance in this context. Objective screening for sleep disturbances in primary care should be prioritized for non-White individuals and men. This is particularly important given that men tend to report fewer sleep problems [12,46] and that both men and racial and ethnic minorities have higher rates of undiagnosed sleep disorders [8,47,48]. Implementing racial and ethnic- and sex-specific programs to monitor and mitigate sleep disparities could also support broader health equity efforts, as poor sleep is a well-established risk factor for various physical and psychological conditions, reduced quality of life, and increased mortality [1–5,49].

## Strengths and Limitations

This study benefits from a diverse, community-based cohort of older adults, including not only NHW and Black participants but also MA individuals with nearly 60% of non-White participants. Additional strengths include objective sleep measures, relatively large sample size, and consideration of numerous important confounders. However, information that could influence sleep disparities, such as occupational history (e.g., night shift work) and residential environment (e.g., urban vs. rural setting, noise, neighborhood safety) was not available in this study. Nonetheless, the potential impact of occupation might be limited given that we controlled for retirement status and 60% of the sample was retired. As this is a cross-sectional study with a single sleep measurement, it does not capture changes in sleep patterns over time. Future longitudinal studies with repeated objective sleep measures across diverse populations are needed to better understand the dynamics and drivers of sleep disparities.

## Conclusions

Our findings reveal significant differences in objective sleep measures across sociodemographic groups, independently of socioeconomic and health factors, with non-White participants and men experiencing poorer sleep. Race and ethnicity and sex were identified as key sociodemographic factors contributing to sleep disparities. These results suggest that poor sleep quality may contribute to health disparities, particularly among vulnerable populations, emphasizing the need for targeted interventions to support those at higher risk.

## Data Availability

Anonymized data within this article will be made available by request from any qualified investigator.

## List of abbreviations

BMI: Body Mass Index
CI: Confidence Interval
FDR: False Discovery Rate
HABS-HD: Health & Aging Brain Study – Health Disparities
MA: Mexican American
NHW: Non-Hispanic White
OR: Odds Ratios
RAPA: Rapid Assessment of Physical Activity
REI: Respiratory Event Index
SD: Standard Deviation
SFI: Sleep Fragmentation Index
WASO: Wake After Sleep Onset

## Declarations

### Ethics approval and consent to participate

All study participants provided written informed consent, and the Dormir Study was approved by the University of North Texas Health Science Center Institutional and University of California San Francisco Review Boards.

### Consent for publication

Not applicable.

### Authors’ contributions

CC conceived and designed the study, analyzed the data, interpreted the study findings, and drafted the manuscript. KLS, YL, and CP interpreted the study findings and critically revised the manuscript. KY conceived and designed the study, interpreted the study findings, and reviewed the manuscript.

## Acknowledgements

The research team thanks the participants of the Dormir study.

