## Supplementary information for "Objective Sleep Quality in Diverse Older Adults: the Importance of Race and Ethnicity and Sex"

eMethods

***Covariates***

Sociodemographic and health characteristics were self-reported through clinical interviews, and included age, sex, race and ethnicity, years of education, retirement status, household income, current smoking status, and medical history of hypertension, diabetes, stroke, heart attack, and depression. Body mass index (BMI) was calculated as the ratio of weight to height (kg/m^2^). Physical activity was measured using the Rapid Assessment of Physical Activity (RAPA) questionnaire and analyzed either as a continuous variable or dichotomized, with participants classified as physically active if their RAPA score was ≥6 [1]. Cognitive diagnoses were determined using an algorithmic decision tree and subsequently confirmed through a consensus review as normal control, mild cognitive impairment (MCI), and dementia [2]. Sleep medication use was assessed through a self-reported item from the Pittsburgh Sleep Quality Index: “During the past month, how often have you taken medicine (prescribed or “over the counter”) to help you sleep?” [3]. Responses were dichotomized into two groups: those who used sleep medications (less than once a week; once or twice a week; and three or more times a week) and those who did not (not during the past month). In a subsample (n=784), participants used the WatchPAT 200 (Itamar Medical Ltd., Caesarea, Israel) during one night for a home sleep apnea test. The respiratory event index (REI) was used to evaluate the presence of moderate to severe sleep apnea (REI ≥15).

eTable 1. Multivariable-adjusted means of objective continuous sleep measures by race and ethnicity (n=824).

|  | Black  (n=190) | MA  (n=282) | NHW  (n=366) |  |  |
| --- | --- | --- | --- | --- | --- |
|  | Adjusted means (95%CI) | Adjusted means (95%CI) | Adjusted means (95%CI) | *P* value | Direction |
| **Sleep duration**, hour | 7.0 (6.7,7.3) | 7.0 (6.7,7.4) | 7.3 (7.0,7.6) | 0.003* | NHW> B,MA |
| **Sleep efficiency**, % ^a^ | 86.0 (84.5,87.4) | 87.0 (85.5,88.5) | 88.8 (87.4,90.1) | <.0001* | NHW> B,MA |
| **WASO**, min ^b^ | 58.2 (50.5,67.0) | 53.6 (46.2,62.1) | 45.5 (39.5,52.5) | <.0001* | NHW< B,MA |
| **SFI**, % | 34.5 (31.8,37.2) | 29.2 (26.5,32.0) | 27.7 (25.0,30.3) | <.0001* | B> MA,NHW |

Abbreviations: CI, confidence interval; MA, Mexican American; NHW, Non-Hispanic White; SFI, sleep fragmentation index; WASO, wake after sleep onset

Models adjusted for race and ethnicity, sex, age, education, retirement status, cognitive impairment, body mass index, physical activity, smoking, sleep medications, and history of hypertension, diabetes, stroke, heart attack, and depression.

^a^ cube transformation; ^b^ log transformation; * results are significant after false discovery rate correction

eTable 2. Multivariable-adjusted associations between objective sleep measures and sociodemographic groups with further adjustment for household income (n=787).

|  | Black *versus* NHW | MA *versus* NHW |  |  | Women *versus*  Men |  |  | ≥65 years *versus*  <65 years |  |
| --- | --- | --- | --- | --- | --- | --- | --- | --- | --- |
|  | OR (95%CI) | OR (95%CI) | *P* value |  | OR (95%CI) | *P* value |  | OR (95%CI) | *P* value |
| **Sleep duration**, hour |  |  | 0.03* |  |  | 0.003* |  |  | 0.90 |
| <6 | 2.64 (1.38,5.04) | 1.56 (0.77,3.18) |  |  | 0.45 (0.27,0.73) |  |  | 1.14 (0.63,2.06) |  |
| [6-8] | 1 | 1 |  |  | 1 |  |  | 1 |  |
| >8 | 0.85 (0.51,1.44) | 0.79 (0.46,1.35) |  |  | 1.11 (0.74,1.68) |  |  | 1.06 (0.67,1.67) |  |
| **Sleep efficiency**, % |  |  | <.001* |  |  | 0.18 |  |  | 0.51 |
| ≥85 | 1 | 1 |  |  | 1 |  |  | 1 |  |
| <85 | 2.55 (1.56,4.16) | 1.84 (1.08,3.14) |  |  | 0.77 (0.52,1.13) |  |  | 0.86 (0.55,1.35) |  |
| **WASO**, min |  |  | 0.002* |  |  | 0.53 |  |  | 0.34 |
| <41.7 | 1 | 1 |  |  | 1 |  |  | 1 |  |
| [41.7-63.5] | 1.85 (1.10,3.10) | 1.48 (0.88,2.50) |  |  | 1.25 (0.84,1.86) |  |  | 1.02 (0.62,1.56) |  |
| >63.5 | 2.92 (1.72,4.96) | 1.72 (0.99,2.99) |  |  | 1.12 (0.74,1.70) |  |  | 1.01 (0.45,1.18) |  |
| **SFI**, % |  |  | <.0001* |  |  | 0.008* |  |  | 0.83 |
| <22.1 | 1 | 1 |  |  | 1 |  |  | 1 |  |
| [22.1-29.9] | 2.73 (1.57,4.76) | 1.10 (0.66,1.84) |  |  | 0.90 (0.60,1.35) |  |  | 0.93 (0.59,1.46) |  |
| >29.9 | 5.37 (3.08,9.36) | 1.37 (0.80,2.36) |  |  | 0.55 (0.36,0.83) |  |  | 1.07 (0.66,1.73) |  |

Abbreviations: CI, confidence interval; MA, Mexican American; NHW, Non-Hispanic White; SFI, sleep fragmentation index; WASO, wake after sleep onset

Models adjusted for race and ethnicity, sex, age, education, retirement status, cognitive impairment, body mass index, physical activity, smoking, sleep medications, and history of hypertension, diabetes, stroke, heart attack, depression, and household income.

* results are significant after false discovery rate correction

eTable 3. Multivariable-adjusted means of objective continuous sleep measures by race and ethnicity with further adjustment for household income (n=787).

|  | Black  (n=190) | MA  (n=282) | NHW  (n=366) |  |  |
| --- | --- | --- | --- | --- | --- |
|  | Adjusted means (95%CI) | Adjusted means (95%CI) | Adjusted means (95%CI) | *P* value | Direction |
| **Sleep duration**, hour | 6.9 (6.6,7.3) | 7.1 (6.7,7.4) | 7.3 (7.0,7.6) | 0.008* | NHW>MA>B |
| **Sleep efficiency**, % ^a^ | 86.4 (84.9,87.8) | 87.4 (85.8,88.8) | 88.9 (87.6,90.3) | <.0001* | NHW>MA,B |
| **WASO**, min ^b^ | 55.8 (48.3,64.5) | 51.7 (44.4,60.3) | 44.4 (38.4,51.3) | <.0001* | NHW< MA,B |
| **SFI**, % | 34.0 (31.2,36.7) | 28.7 (25.8,31.6) | 27.3 (24.6,30.0) | <.0001* | B> NHW,MA |

Abbreviations: CI, confidence interval; MA, Mexican American; NHW, Non-Hispanic White; SFI, sleep fragmentation index; WASO, wake after sleep onset

Models adjusted for race and ethnicity, sex, age, education, retirement status, cognitive impairment, body mass index, physical activity, smoking, sleep medications, and history of hypertension, diabetes, stroke, heart attack, depression, and household income.

^a^ cube transformation; ^b^ log transformation; * results are significant after false discovery rate correction

eTable 4. Multivariable-adjusted associations between objective sleep measures and sociodemographic groups with further adjustment for moderate to severe sleep apnea (n=784).

|  | Black *versus* NHW | MA *versus* NHW |  |  | Women *versus*  Men |  |  | ≥65 years *versus*  <65 years |  |
| --- | --- | --- | --- | --- | --- | --- | --- | --- | --- |
|  | OR (95%CI) | OR (95%CI) | *P* value |  | OR (95%CI) | *P* value |  | OR (95%CI) | *P* value |
| **Sleep duration**, hour |  |  | 0.04* |  |  | 0.002* |  |  | 0.97 |
| <6 | 2.31 (1.19,4.48) | 1.41 (0.69,2.89) |  |  | 0.46 (0.28,0.77) |  |  | 0.97 (0.52,1.80) |  |
| [6-8] | 1 | 1 |  |  | 1 |  |  | 1 |  |
| >8 | 0.76 (0.45,1.28) | 0.67 (0.40,1.14) |  |  | 1.30 (0.86,1.95) |  |  | 1.05 (0.66,1.67) |  |
| **Sleep efficiency**, % |  |  | 0.005* |  |  | 0.05 |  |  | 0.28 |
| ≥85 | 1 | 1 |  |  | 1 |  |  | 1 |  |
| <85 | 2.28 (1.38,3.76) | 1.68 (0.99,2.85) |  |  | 0.68 (0.46,1.00) |  |  | 0.78 (0.49,1.23) |  |
| **WASO**, min |  |  | 0.004* |  |  | 0.31 |  |  | 0.22 |
| <41.7 | 1 | 1 |  |  | 1 |  |  | 1 |  |
| [41.7-63.5] | 1.96 (1.17,3.30) | 1.66 (1.00,2.77) |  |  | 1.35 (0.91,2.00) |  |  | 0.94 (0.60,1.50) |  |
| >63.5 | 2.67 (1.56,4.58) | 1.78 (1.04,3.05) |  |  | 1.11 (0.74,1.67) |  |  | 0.68 (0.42,1.10) |  |
| **SFI**, % |  |  | <.0001* |  |  | 0.003* |  |  | 0.91 |
| <22.1 | 1 | 1 |  |  | 1 |  |  | 1 |  |
| [22.1-29.9] | 2.56 (1.46,4.50) | 0.90 (0.55,1.50) |  |  | 0.94 (0.63,1.41) |  |  | 1.10 (0.70,1.75) |  |
| >29.9 | 4.77 (2.71,8.39) | 1.31 (0.77,2.22) |  |  | 0.53 (0.35,0.79) |  |  | 1.05 (0.65,1.71) |  |

Abbreviations: CI, confidence interval; MA, Mexican American; NHW, Non-Hispanic White; SFI, sleep fragmentation index; WASO, wake after sleep onset

Models adjusted for race and ethnicity, sex, age, education, retirement status, cognitive impairment, body mass index, physical activity, smoking, sleep medications, and history of hypertension, diabetes, stroke, heart attack, depression, and moderate to severe sleep apnea.

* results are significant after false discovery rate correction

eTable 5. Multivariable-adjusted means of objective continuous sleep measures by race and ethnicity with further adjustment for moderate to severe sleep apnea (n=784).

|  | Black  (n=190) | MA  (n=282) | NHW  (n=366) |  |  |
| --- | --- | --- | --- | --- | --- |
|  | Adjusted means (95%CI) | Adjusted means (95%CI) | Adjusted means (95%CI) | *P* value | Direction |
| **Sleep duration**, hour | 6.9 (6.6,7.2) | 6.9 (6.6,7.3) | 7.2 (6.9,7.5) | 0.006* | NHW> MA,B |
| **Sleep efficiency**, % ^a^ | 86.1 (84.6,87.6) | 86.9 (85.4,88.4) | 88.6 (87.2,90.0) | <.0001* | NHW> MA,B |
| **WASO**, min ^b^ | 57.3 (49.4,66.4) | 53.4 (45.9,62.1) | 45.8 (39.6,53.1) | <.0001* | NHW< MA,B |
| **SFI**, % | 34.7 (31.9,37.4) | 29.6 (26.8,32.5) | 28.3 (25.5,31.0) | <.0001* | B> NHW,MA |

Abbreviations: CI, confidence interval; MA, Mexican American; NHW, Non-Hispanic White; SFI, sleep fragmentation index; WASO, wake after sleep onset

Models adjusted for race and ethnicity, sex, age, education, retirement status, cognitive impairment, body mass index, physical activity, smoking, sleep medications, and history of hypertension, diabetes, stroke, heart attack, depression, and moderate to severe sleep apnea.

^a^ cube transformation; ^b^ log transformation; * results are significant after false discovery rate correction

eTable 6. Multivariable-adjusted means of objective continuous sleep measures by sex and age groups (n=824).

|  | Men  (n=290) | Women  (n=534) |  |  | <65 years  (n=340) | ≥65 years  (n=484) |  |
| --- | --- | --- | --- | --- | --- | --- | --- |
|  | Adjusted means (95% CI) | Adjusted means (95% CI) | *P* value |  | Adjusted means (95% CI) | Adjusted means (95% CI) | *P* value |
| **Sleep duration**, hour | 7.0 (6.7,7.2) | 7.3 (7.0,7.6) | 0.0002* |  | 7.0 (6.8,7.3) | 7.2 (6.9,7.5) | 0.22 |
| **Sleep efficiency**, % ^a^ | 86.9 (85.5,88.2) | 87.7 (86.3,89.0) | 0.03* |  | 87.0 (85.6,88.3) | 87.6 (86.2,88.9) | 0.18 |
| **WASO**, min ^b^ | 52.2 (45.5,59.9) | 52.1 (45.5,59.7) | 0.98 |  | 53.8 (46.9,61.8) | 50.6 (44.0,58.1) | 0.18 |
| **SFI**, % | 32.2 (29.6,34.8) | 28.7 (26.2,31.3) | <.0001* |  | 30.5 (27.9,33.1) | 30.5 (27.8,33.1) | 0.99 |

Abbreviations: CI, confidence interval; SFI, sleep fragmentation index; WASO, wake after sleep onset

Models adjusted for race and ethnicity, sex, age, education, retirement status, cognitive impairment, body mass index, physical activity, smoking, sleep medications, and history of hypertension, diabetes, stroke, heart attack, and depression.

^a^ cube transformation; ^b^ log transformation; * results are significant after false discovery rate correction

eTable 7. Multivariable-adjusted means of objective continuous sleep measures by sex and age groups with further adjustment for household income (n=787).

|  | Men  (n=290) | Women  (n=534) |  |  | <65 years  (n=340) | ≥65 years  (n=484) |  |
| --- | --- | --- | --- | --- | --- | --- | --- |
|  | Adjusted means (95% CI) | Adjusted means (95% CI) | *P* value |  | Adjusted means (95% CI) | Adjusted means (95% CI) | *P* value |
| **Sleep duration**, hour | 7.0 (6.7,7.3) | 7.2 (6.9,7.5) | 0.002* |  | 7.0 (6.7,7.4) | 7.1 (6.8,7.4) | 0.37 |
| **Sleep efficiency**, % ^a^ | 87.2 (85.8,88.6) | 87.9 (86.5,89.3) | 0.07 |  | 87.3 (85.8,88.6) | 87.9 (86.5,89.2) | 0.16 |
| **WASO**, min ^b^ | 50.4 (43.8,57.9) | 50.5 (43.8,58.1) | 0.97 |  | 52.2 (45.4,60.1) | 48.7 (42.2,56.1) | 0.13 |
| **SFI**, % | 31.7 (29.1,34.3) | 28.2 (25.6,30.9) | <.0001* |  | 30.0 (27.3,32.6) | 30.0 (27.3,32.7) | 0.95 |

Abbreviations: CI, confidence interval; SFI, sleep fragmentation index; WASO, wake after sleep onset

Models adjusted for race and ethnicity, sex, age, education, retirement status, cognitive impairment, body mass index, physical activity, smoking, sleep medications, and history of hypertension, diabetes, stroke, heart attack, depression, and household income.

^a^ cube transformation; ^b^ log transformation; * results are significant after false discovery rate correction

eTable 8. Multivariable-adjusted means of objective continuous sleep measures by sex and age groups with further adjustment for moderate to severe sleep apnea (n=784).

|  | Men  (n=290) | Women  (n=534) |  |  | <65 years  (n=340) | ≥65 years  (n=484) |  |
| --- | --- | --- | --- | --- | --- | --- | --- |
|  | Adjusted means (95% CI) | Adjusted means (95% CI) | *P* value |  | Adjusted means (95% CI) | Adjusted means (95% CI) | *P* value |
| **Sleep duration**, hour | 6.9 (6.6,7.2) | 7.2 (6.9,7.5) | 0.0003* |  | 7.0 (6.7,7.2) | 7.1 (6.8,7.4) | 0.34 |
| **Sleep efficiency**, % ^a^ | 86.8 (85.4,88.2) | 87.6 (86.3,89.0) | 0.04 |  | 86.9 (85.4,88.2) | 87.6 (86.2,89.0) | 0.10 |
| **WASO**, min ^b^ | 51.9 (45.0,59.9) | 51.9 (45.2,59.8) | 0.99 |  | 54.0 (46.7,62.3) | 50.0 (43.3,57.7) | 0.10 |
| **SFI**, % | 32.6 (29.9,35.3) | 29.1 (26.5,31.7) | <.0001* |  | 30.9 (28.2,33.6) | 30.7 (28.0,33.4) | 0.84 |

Abbreviations: CI, confidence interval; SFI, sleep fragmentation index; WASO, wake after sleep onset

Models adjusted for race and ethnicity, sex, age, education, retirement status, cognitive impairment, body mass index, physical activity, smoking, sleep medications, and history of hypertension, diabetes, stroke, heart attack, depression, and moderate to severe sleep apnea.

^a^ cube transformation; ^b^ log transformation; * results are significant after false discovery rate correction

eTable 9. Objective categorical sleep measures by race and ethnicity and sex.

|  | Men  (n=295) | | | |  | Women  (n=543) | | | |
| --- | --- | --- | --- | --- | --- | --- | --- | --- | --- |
|  | Black  (n=59) | MA  (n=84) | NHW  (n=152) |  |  | Black  (n=131) | MA  (n=198) | NHW  (n=214) |  |
|  | No. (%) | No. (%) | No. (%) | *P* value^a^ |  | No. (%) | No. (%) | No. (%) | *P* value^a^ |
| **Sleep duration**, hour |  |  |  | 0.0008* |  |  |  |  | 0.002* |
| <6 | 18 (30.5) | 19 (22.6) | 14 (9.2) |  |  | 20 (15.3) | 23 (11.6) | 9 (4.2) |  |
| [6-8] | 31 (52.5) | 55 (65.5) | 101 (66.4) |  |  | 84 (64.1) | 131 (66.2) | 137 (64.0) |  |
| >8 | 10 (16.9) | 10 (11.9) | 37 (24.3) |  |  | 27 (20.6) | 44 (22.2) | 68 (31.8) |  |
| **Sleep efficiency**, % |  |  |  | <.0001* |  |  |  |  | <.0001* |
| ≥85 | 32 (54.2) | 52 (61.9) | 129 (84.9) |  |  | 92 (70.2) | 140 (70.7) | 189 (88.3) |  |
| <85 | 27 (45.8) | 32 (38.1) | 23 (15.1) |  |  | 39 (29.8) | 58 (29.3) | 25 (11.7) |  |
| **WASO**, min |  |  |  | <.0001* |  |  |  |  | <.0001* |
| <41.4 | 8 (13.6) | 25 (29.8) | 77 (50.7) |  |  | 33 (25.2) | 43 (21.7) | 93 (43.5) |  |
| [41.4-63.7] | 20 (33.9) | 26 (31.0) | 41 (27.0) |  |  | 44 (33.6) | 73 (36.9) | 76 (35.5) |  |
| >63.7 | 31 (52.5) | 33 (39.3) | 34 (22.4) |  |  | 54 (41.2) | 82 (41.4) | 45 (21.0) |  |
| **SFI**,% |  |  |  | <.0001* |  |  |  |  | <.0001* |
| <22.1 | 3 (5.1) | 24 (28.6) | 65 (42.8) |  |  | 25 (19.1) | 64 (32.3) | 99 (46.3) |  |
| [22.1-29.9] | 20 (33.9) | 18 (21.4) | 46 (30.3) |  |  | 42 (32.1) | 79 (39.9) | 73 (34.1) |  |
| >29.9 | 36 (61.0) | 42 (50.0) | 41 (27.0) |  |  | 64 (48.9) | 55 (27.8) | 42 (19.6) |  |

Abbreviations: MA, Mexican American; NHW, Non-Hispanic White; SFI, sleep fragmentation index; WASO, wake after sleep onset

^a^ Chi-square test was used
* results are significant after false discovery rate correction

eTable 10. Description of covariates by sex and age groups.

|  | Men  (n=295) | Women  (n=543) |  |  | <65 years  (n=346) | ≥65 years  (n=492) |  |
| --- | --- | --- | --- | --- | --- | --- | --- |
|  | No. (%) or mean (±SD) | No. (%) or mean (±SD) | *P* value^b^ |  | No. (%) or mean (±SD) | No. (%) or mean (±SD) | *P* value^b^ |
| Race and Ethnicity |  |  | 0.003 |  |  |  | <.0001 |
| Black | 59 (20.0) | 131 (24.1) |  |  | 107 (30.9) | 83 (16.9) |  |
| MA | 84 (28.5) | 198 (36.5) |  |  | 146 (42.2) | 136 (27.6) |  |
| NHW | 152 (51.5) | 214 (39.4) |  |  | 93 (26.9) | 273 (55.5) |  |
| Sex, *women* | - | - | - |  | 244 (70.5) | 299 (60.8) | 0.004 |
| Age, years | 68.4 (±8.5) | 65.9 (±8.2) | <.0001 |  | 58.7 (±4.0) | 72.4 (±5.7) | <.0001 |
| Age, *≥65 years* | 193 (65.4) | 299 (55.1) | 0.004 |  | - | - | - |
| Education, years | 14.3 (±4.0) | 13.5 (±4.0) | 0.005 |  | 13.3 (±4.0) | 14.2 (±4.0) | 0.003 |
| Retirement status | 190 (64.6) | 317 (58.5) | 0.08 |  | 91 (26.4) | 416 (84.7) | <.0001 |
| Income groups |  |  | 0.001 |  |  |  | 0.57 |
| <25,000 | 60 (21.2) | 155 (30.0) |  |  | 92 (27.5) | 123 (26.4) |  |
| [25,000-50,000[ | 58 (20.5) | 135 (26.1) |  |  | 80 (24.0) | 113 (24.2) |  |
| [50,000-75,000[ | 55 (19.4) | 69 (13.3) |  |  | 45 (13.5) | 79 (17.0) |  |
| ≥75,000 | 110 (38.9) | 158 (30.6) |  |  | 117 (35.0) | 151 (32.4) |  |
| Cognitive impairment | 94 (31.9) | 94 (17.3) | <.0001 |  | 80 (23.1) | 108 (22.0) | 0.69 |
| Smoke currently | 26 (8.8) | 24 (4.4) | 0.01 |  | 27 (7.8) | 23 (4.7) | 0.06 |
| RAPA score ^a^ | 4 (3,6) | 4 (3,6) | 0.002 |  | 4 (3,6) | 4 (3,6) | 0.17 |
| BMI, kg/m^2^ ^a^ | 29.6 (26.3,33.5) | 30.1 (26.1,35.1) | 0.63 |  | 30.8 (26.7,36.6) | 29.3 (25.7,33.1) | <.0001 |
| Sleep medications | 101 (34.2) | 206 (37.9) | 0.29 |  | 130 (37.6) | 177 (36.0) | 0.64 |
| History of hypertension | 207 (70.2) | 330 (60.8) | 0.007 |  | 202 (58.4) | 335 (68.1) | 0.004 |
| History of diabetes | 82 (27.8) | 128 (23.6) | 0.18 |  | 89 (25.7) | 121 (24.6) | 0.71 |
| History of stroke | 13 (4.4) | 29 (5.3) | 0.55 |  | 13 (3.8) | 29 (5.9) | 0.16 |
| History of heart attack | 15 (5.1) | 18 (3.3) | 0.21 |  | 7 (2.0) | 26 (5.3) | 0.02 |
| History of depression | 75 (25.4) | 215 (39.6) | <.0001 |  | 131 (37.9) | 159 (32.3) | 0.10 |
| REI, *≥15* | 139 (51.1) | 260 (50.8) | 0.93 |  | 163 (50.0) | 236 (51.5) | 0.67 |

Abbreviations: BMI, body mass index; MA, Mexican American; NHW, Non-Hispanic White; RAPA, rapid assessment of physical activity; REI, respiratory event index; SD, standard deviation
^a^ median (interquartile range) ^b^ T-test was used for continuous variables with a normal distribution, Mann-Whitney U test for continuous variables without a normal distribution and, Chi-square test for categorical variables

eFigure 1. Stepwise approach to identify most important contributors in the association between sleep duration and age groups.

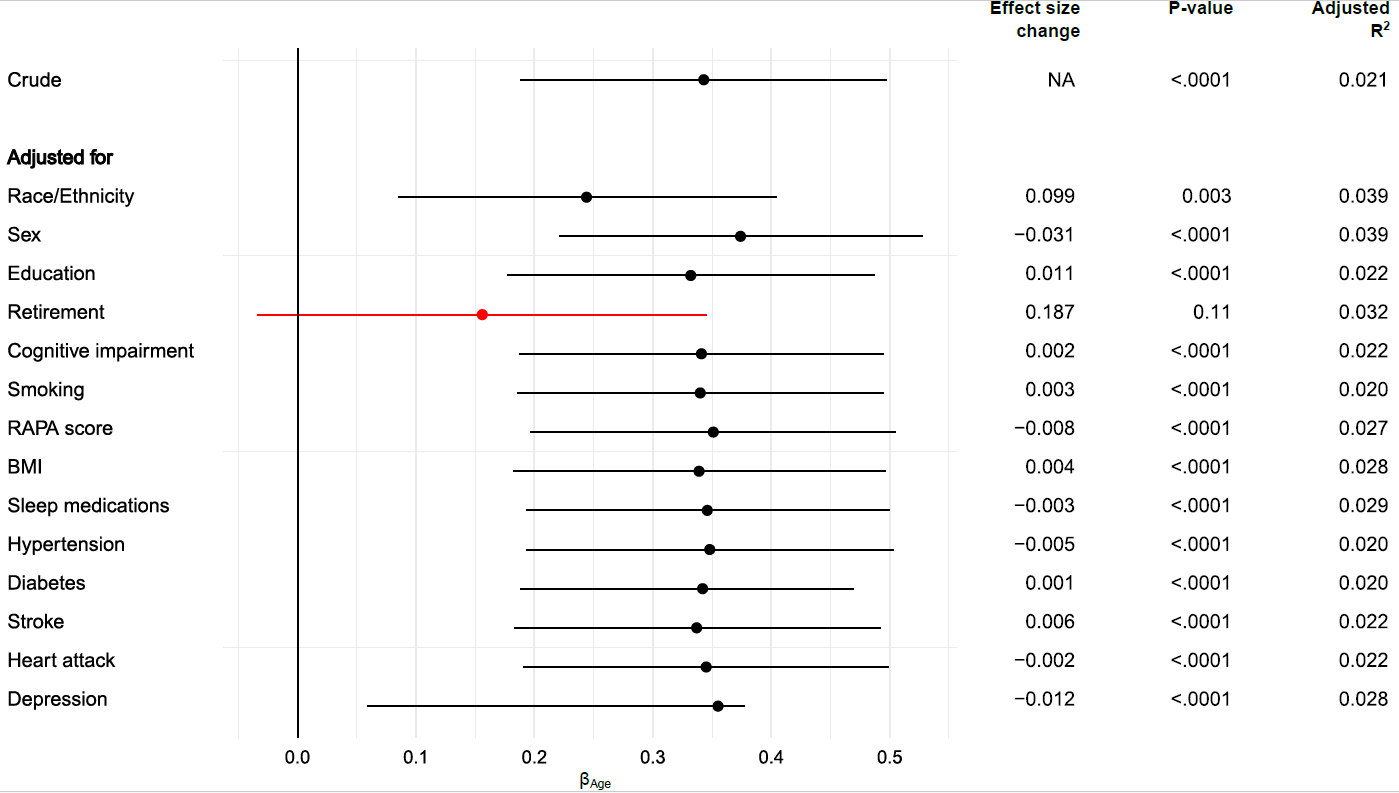

Abbreviations: BMI, body mass index; NA, not applicable; RAPA, rapid assessment of physical activity

Effect size change corresponds to the subtraction of the beta associated with the variable age in the crude model with the beta associated with age in each adjusted model.

eFigure 2. Stepwise approach to identify most important contributors in the association between sleep efficiency and age groups.

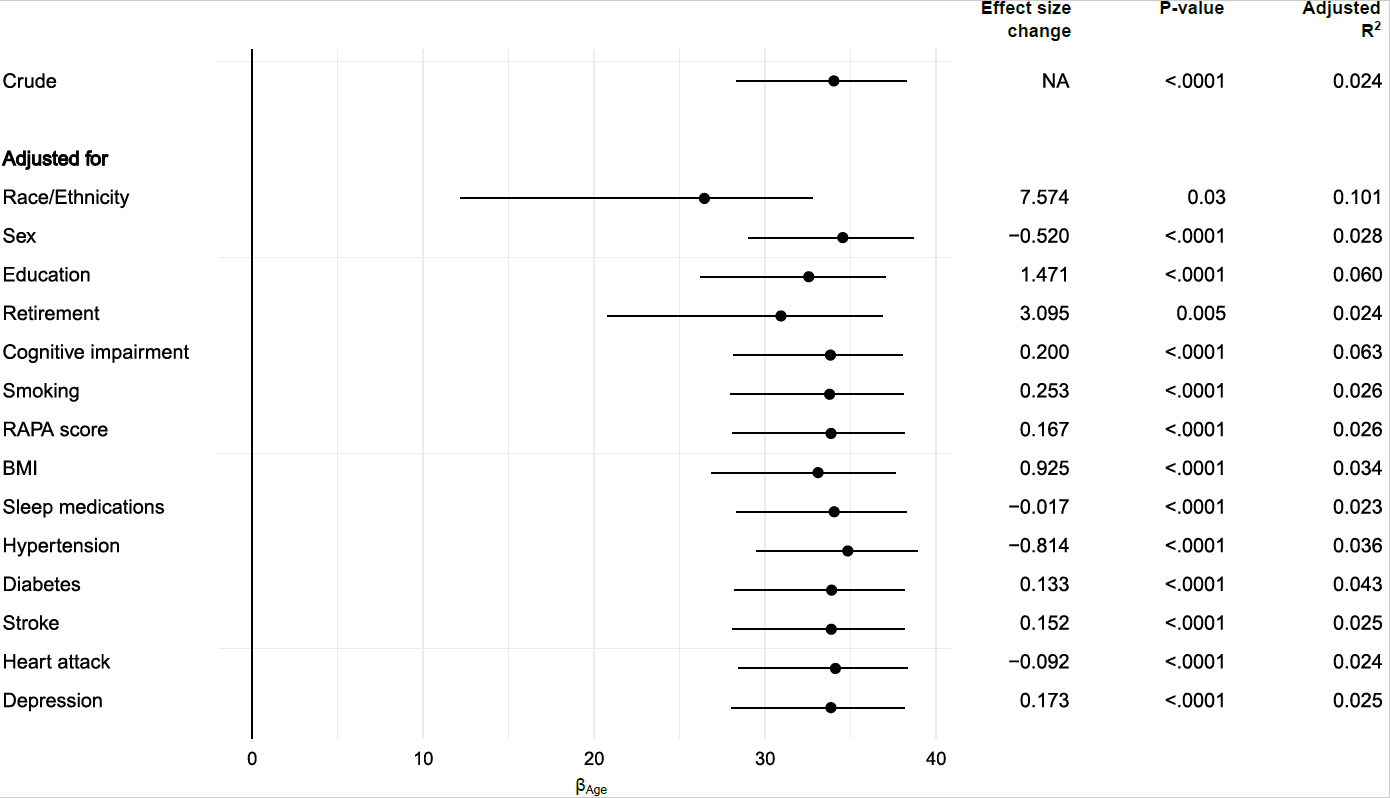

Abbreviations: BMI, body mass index; NA, not applicable; RAPA, rapid assessment of physical activity

Effect size change corresponds to the subtraction of the beta associated with the variable age in the crude model with the beta associated with age in each adjusted model.

eFigure 3. Stepwise approach to identify most important contributors in the association between wake after sleep onset and age groups.

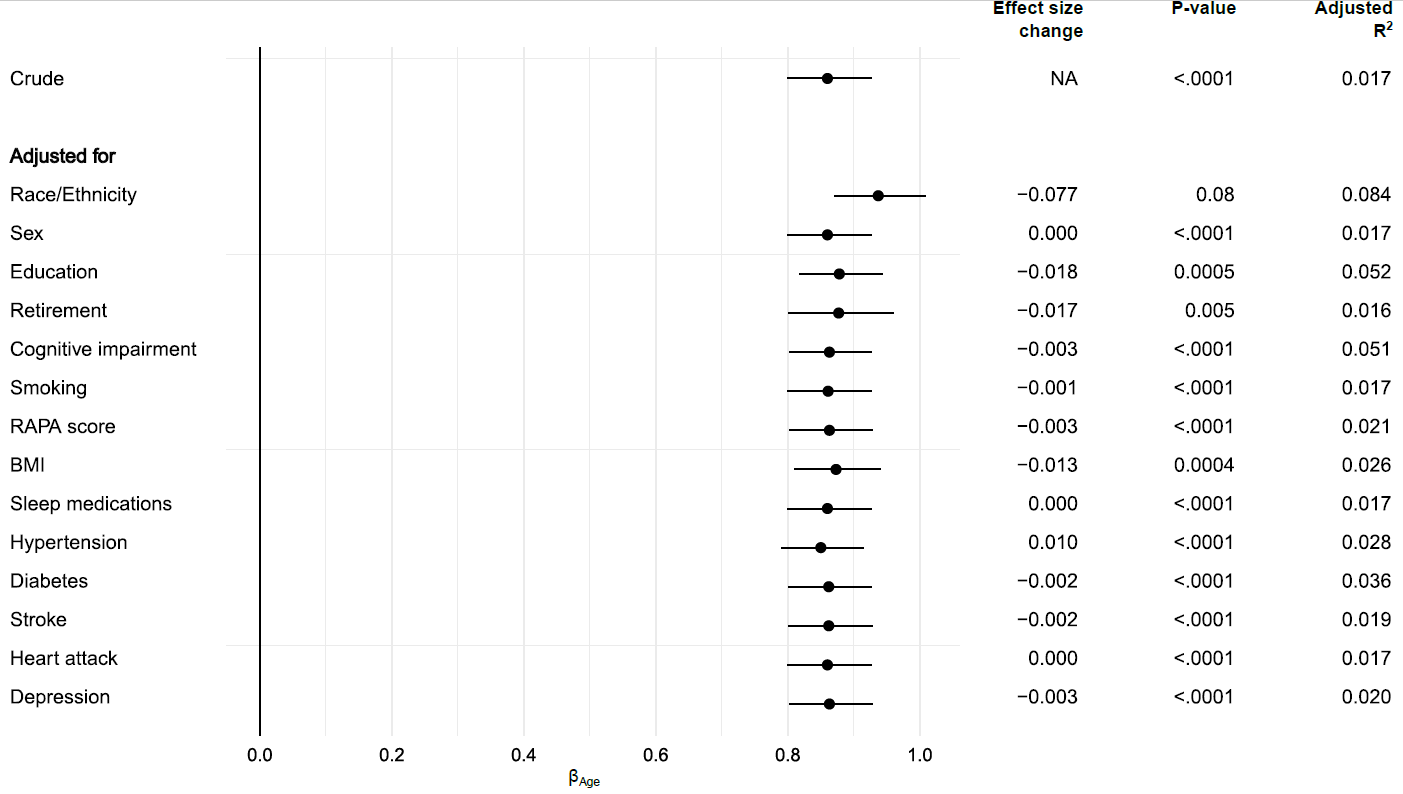

Abbreviations: BMI, body mass index; NA, not applicable; RAPA, rapid assessment of physical activity

Effect size change corresponds to the subtraction of the beta associated with the variable age in the crude model with the beta associated with age in each adjusted model.

eFigure 4. Stepwise approach to identify most important contributors in the association between sleep fragmentation index and age groups.

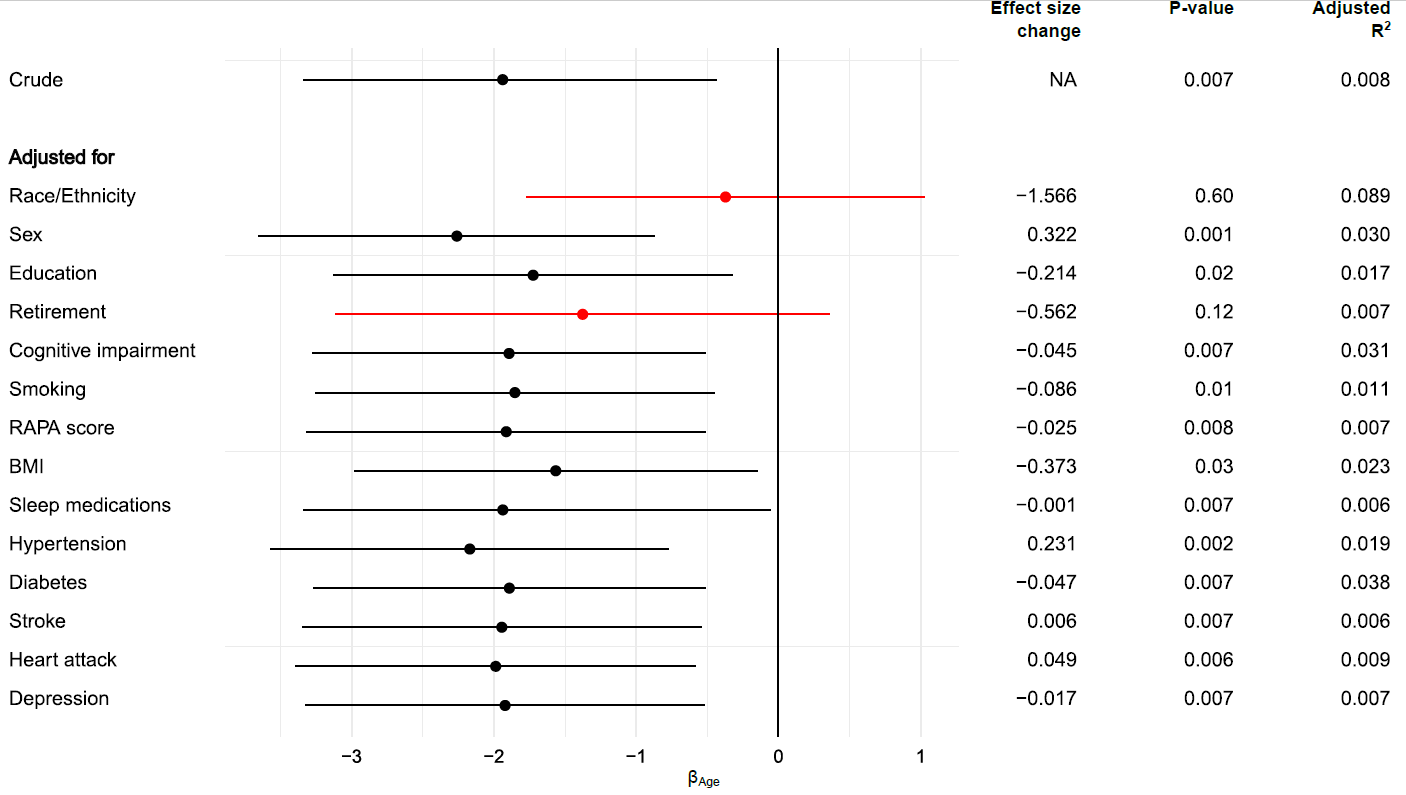

Abbreviations: BMI, body mass index; NA, not applicable; RAPA, rapid assessment of physical activity

Effect size change corresponds to the subtraction of the beta associated with the variable age in the crude model with the beta associated with age in each adjusted model.
